# Ebola virus transmission among healthcare workers: genomic epidemiological analysis of the 2014 Kenema Government Hospital outbreak

**DOI:** 10.64898/2026.10.26.26366557

**Authors:** Christopher Y. Shin, Mambu Momoh, Christopher Tomkins-Tinch, Augustine Goba, Lansana Kanneh, Foday Alhassan, Michael Gbakie, Mohamed Alex Vandi, Ivan Specht, Patrick Varilly, John S. Schieffelin, Shirlee Wohl, Danny Park, Kristian G. Andersen, Donald Grant, Robert F. Garry, Kayla Barnes, Pardis C. Sabeti

**Affiliations:** Harvard College, Harvard University, Cambridge, MA, USA; Broad Institute of MIT and Harvard, Cambridge, MA, USA; Biomedical Sciences Graduate Program, Department of Microbiology and Immunology, Tulane University School of Medicine, New Orleans, LA, USA; Kenema Government Hospital, Ministry of Health and Sanitation, Kenema, Sierra Leone; Eastern Technical University of Sierra Leone, Kenema, Sierra Leone; College of Medicine and Allied Health Sciences, University of Sierra Leone, Freetown, Sierra Leone; National Public Health Agency, Freetown, Sierra Leone; Institute for Computational and Mathematical Engineering, School of Engineering, Stanford University, Stanford, CA, USA; Department of Pediatrics, Tulane University School of Medicine, New Orleans, LA, United States; Division of Infectious Diseases, Brigham and Women’s Hospital, Boston, USA; Department of Immunology and Microbiology, The Scripps Research Institute, La Jolla, CA, USA; Scripps Research Translational Institute, La Jolla, CA, USA; Department of Clinical Sciences, Liverpool School of Tropical Medicine, Liverpool, UK; Department of Microbiology and Immunology, Tulane University Health Sciences Center, New Orleans, LA, USA; Malawi Liverpool Wellcome Research Programme, Blantyre, Malawi; Vector Department, Liverpool School of Tropical Medicine, Liverpool, UK; Department of Organismic and Evolutionary Biology, Harvard University, Cambridge, MA, USA; Harvard Medical School, Boston, MA, USA; Department of Immunology and Infectious Disease, Harvard T.H. Chan School of Public Health, Boston, MA, USA; Howard Hughes Medical Institute, Chevy Chase, MD, USA

## Abstract

**Background:** During the 2014 Ebola outbreak in Sierra Leone, widespread infections among healthcare workers at Kenema Government Hospital severely disrupted clinical operations. Whether these infections primarily arose from repeated patient exposures or from transmission among colleagues remained unclear, particularly as patient admissions exceeded the hospital’s capacity for safe isolation and care.

**Methods:** We revisited the outbreak using expanded viral surveillance and classical genomic analysis, Bayesian transmission inference with *juniper*, and firsthand epidemiologic accounts from hospital staff. We analyzed 279 Ebola virus genomes from Kenema District, including 35 from infected healthcare workers, and 514 genomes in the broader transmission reconstruction, explicitly accounting for unsampled cases.

**Results:** The 35 healthcare worker genomes comprised 28 haplotypes, including five shared by multiple cases. In the reconstructed transmission network, 22 candidate healthcare worker-to-healthcare worker links exceeded a posterior probability threshold of 5%, representing 6.1-fold enrichment over expectation. Healthcare workers accounted for 33.2% of expected onward transmissions at their peak in the two-week window beginning June 23, 2014. Firsthand anecdotal accounts situated inferred relationships within specific wards, clinical procedures, and working relationships of KGH, including care of infected colleagues and infections among laboratory, pharmacy, and other personnel beyond routine bedside care.

**Conclusions:** Ebola transmission involved interconnected groups of healthcare workers across multiple hospital departments during a period of severe resource constraints and expanding patient demand. These findings underscore the importance of protecting and surveilling healthcare workers as an interconnected workforce during emerging outbreaks.

## Introduction

Healthcare workers (HCWs) are among the populations at greatest risk during outbreaks of viral hemorrhagic fever^1–3^. They care for highly infectious patients, often under conditions of resource scarcity and intense operational strain, and have experienced disproportionate infection and mortality during outbreaks of Ebola virus disease (EVD), Marburg virus disease, and Lassa fever^3–10^. These infections not only cost lives but can erode the workforce needed to sustain an outbreak response^4^. Protecting HCWs is therefore essential both for individual safety and for maintaining the functioning of the health system.

The 2013–2016 West African EVD epidemic, the largest EVD outbreak recorded to date, exposed the consequences of healthcare-associated transmission at unprecedented scale^11^. As Ebola spread across Guinea, Sierra Leone, and Liberia, healthcare facilities became major sites of exposure, and frontline workers faced repeated risk through patient care and within health systems under rapidly escalating pressure^4,6,11–13^.

Kenema Government Hospital (KGH) was at the center of Sierra Leone’s early response^6,14^. For decades, KGH had served as a regional referral center for Lassa fever, with specialized clinical and laboratory capacity for viral hemorrhagic fever diagnosis and care^6,14,15^. After EVD was confirmed in Guinea, the KGH Lassa fever team rapidly established Ebola diagnostic capacity, confirmed Sierra Leone’s first Ebola case, and converted the Lassa fever ward into the country’s first dedicated Ebola treatment unit as cases increased^6,12,15,16^. KGH’s deep expertise with viral hemorrhagic fever could not compensate for the lack of protective infrastructure needed for an outbreak of this scale. During the early months of the epidemic, shortages of personal protective equipment (PPE), clean water, chlorine, reliable electricity, trained personnel, and isolation capacity constrained infection prevention and control^1,6,15^. Clinical and laboratory staff at times cared for large numbers of highly infectious patients with limited resources, while infections among healthcare workers further reduced staffing and intensified pressure on the response^4,6^.

Although HCW infections became a defining feature of the 2014–2016 West African EVD epidemic, it remained unclear how those infections arose and spread. Infection prevention appropriately focuses on exposure from infected patients, but healthcare workers are also part of a highly interconnected workforce, interacting across wards, procedures, handoffs, shared spaces, and care of sick colleagues.^6^ At KGH, where the workforce was heavily affected, a central unresolved question was whether infections primarily reflected separate exposures to infected patients or whether Ebola had also begun to propagate among healthcare workers themselves.

Reconstructing these transmission pathways has historically been difficult. Outbreaks often yield too few sampled cases, incomplete epidemiologic information, or insufficient genomic coverage to resolve transmission pathways. Even when dense sequencing is available, viral genomes can change too slowly relative to the pace of transmission to distinguish closely related infections: during the West African Ebola epidemic, successive cases could carry identical or nearly identical consensus genomes, leaving both the direction of transmission and the possibility of unsampled intermediates unresolved^12,17,18^. More recently, transmission-reconstruction approaches that jointly model phylogeny, transmission, and unsampled cases have made it possible to extract substantially more information from densely sampled outbreak datasets^19,20^.

Here, we revisit the KGH outbreak amongst HCWs using expanded viral genomic sequencing, transmission inference with juniper, epidemiologic investigation, and firsthand accounts from KGH staff. We sought to assess the evidence for transmission among healthcare workers, examine plausible pathways across wards, clinical events, and working relationships, and understand how these interactions may have heightened the vulnerability of the hospital workforce.

## Results

### Classical genomic analysis identifies a closely related HCW cluster but cannot resolve transmission

To reconstruct transmission among healthcare workers at KGH, we expanded existing genomic surveillance to capture substantially more of the infected healthcare workforce. Earlier sequencing efforts established that the Sierra Leone epidemic descended from a single introduction across the Guinean border and documented sustained local transmission within Kenema District, but were not designed to examine transmission within the KGH healthcare workforce^12,18^. We therefore generated 63 additional Ebola virus genomes from KGH patients and staff, including 21 previously unsequenced healthcare workers. Together with our earlier sequencing efforts, this yielded a dataset comprising 279 of 591 (47%) confirmed EVD cases reported in Kenema District during the May–November 2014 period and 35 of 79 infected KGH healthcare workers during that period (44%)^6^.

Expanded genomic sampling revealed that healthcare worker infections were concentrated in several closely related clusters within the broader Kenema outbreak. Using IQ-TREE, we inferred a maximum-likelihood phylogeny from the Kenema District consensus genomes, with branch lengths representing substitutions per site (Figure 1A)^21^. HCW sequences were concentrated within a limited region of the phylogeny constructed from cases sequenced at KGH, many of which originating in Kenema District, while other HCW sequences were interspersed among non-Kenema District cases. Because KGH also cared for patients referred from other districts, this phylogeny does not represent all patients treated at KGH. HCW infections were genomically concentrated but did not form a single monophyletic clade.

**Figure 1.**
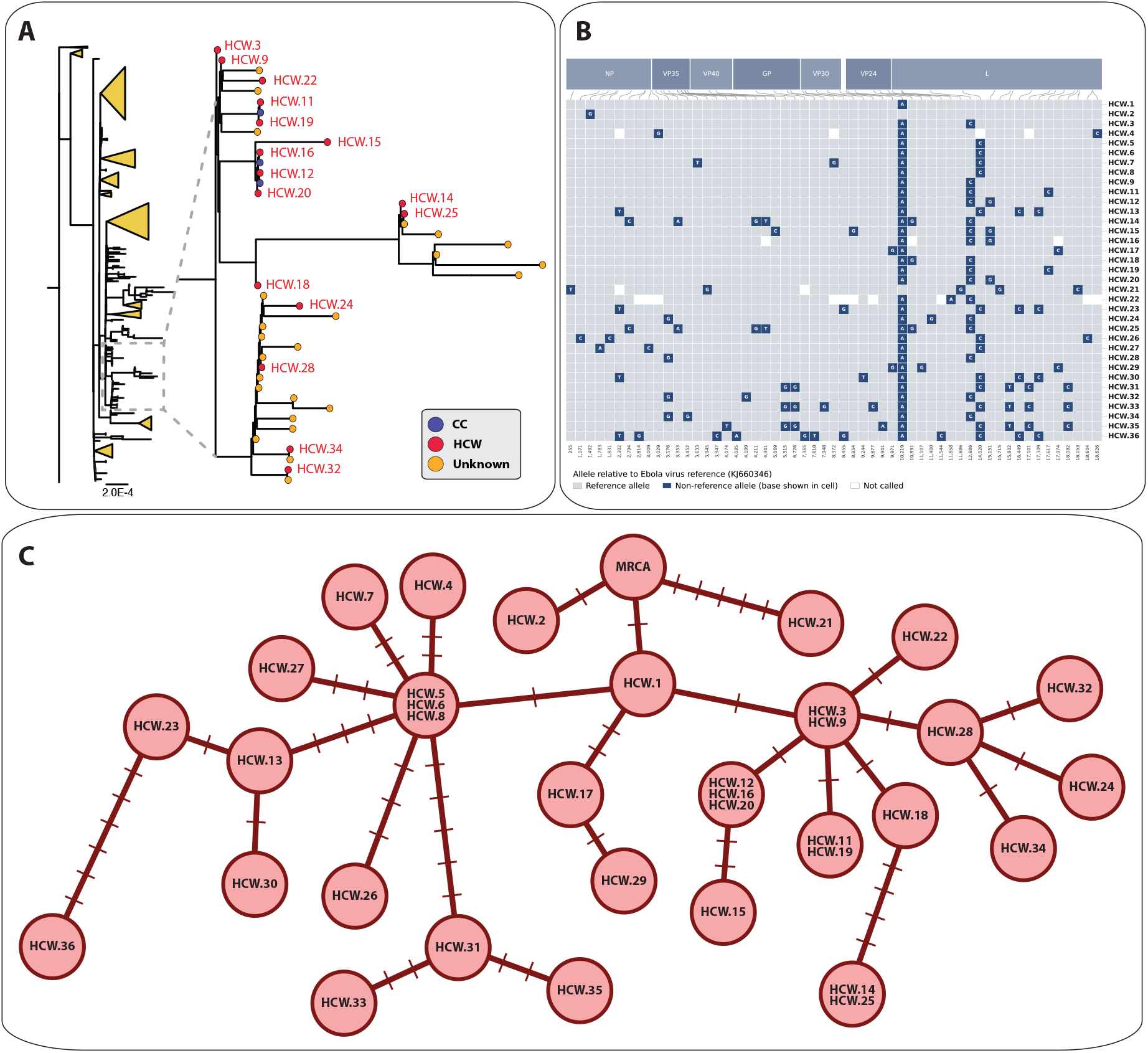
Classical genomic analysis of Ebola virus genome sequences from Kenema Government Hospital. **(A)** Maximum-likelihood phylogeny of 279 Ebola virus genomes from Kenema District, with patient-only clades collapsed into triangles for display. Inset: expanded view of the region of the tree containing HCW sequences. Tips are colored by case status (red, healthcare worker (HCW); blue, close contact of healthcare worker (CC); yellow, patient with unknown HCW status), with HCW cases labeled in red. Scale bar indicates substitutions per site. **(B)** Consensus single-nucleotide variant matrix for the 35 HCW genomes, restricted to positions variable within the cohort and ordered by genomic coordinate. Light cells indicate the reference allele, dark cells a non-reference allele, and gray cells positions with no unambiguous call. Gene products are indicated above; genomic positions are labeled at top and case identifiers at right. ADAR-masked sites are excluded. **(C)** Haplotype network of the 35 HCW consensus genomes. Sequences with identical consensus genotypes are collapsed into a single node, with node size scaled to the number of sequences it contains; nodes are joined by a minimum spanning tree over pairwise SNP distance. Each tick mark on an edge denotes one single-nucleotide difference, labeled by its position in the KJ660346.2 reference genome coordinate system.

Genomic comparison showed that many HCW infections were separated by very little sequence variation. The 35 sequences collapsed into 28 distinct haplotypes, with most neighboring haplotypes separated by only one or two substitutions and five haplotypes shared by more than one case (Figure 1C). The network was consistent with an infinite-sites model, with each of the 55 variable consensus sites (Figure 1B) represented by exactly one substitution across the network (Figure 1C), requiring no recurrent mutations or reversions. Several epidemiologically proximate HCWs carried identical consensus genomes, while others differed by only one or two substitutions.

The evolutionary rate of Ebola virus helps explain why classical genomic analysis has limited resolution among closely spaced transmission events. Published estimates from the West African epidemic place the evolutionary rate at approximately 1.2 × 10−^3^ substitutions per site per year (95% CrI: 1.13–1.27 × 10−^3^ ), corresponding to roughly one consensus substitution across the 18.9-kb genome every 16 days^22,23^. With mean serial intervals of approximately 14–15 days, successive infections could carry identical consensus genomes, while differences of one or two substitutions could not distinguish direct transmission from chains involving unsampled intermediates^24^. Classical genomic analysis thus established a closely related and clustered set of HCW infections at KGH, but could not determine the direction or probability of transmission among them.

### Transmission inference reveals a densely interconnected healthcare worker network at KGH

To reconstruct likely transmission pathways among closely related cases, we applied *juniper*, a Bayesian framework that jointly infers phylogeny and transmission while modeling unsampled cases^19^. For this analysis, we expanded the dataset beyond the 279 Kenema District genomes to include 235 additional Ebola virus genomes sampled elsewhere in Sierra Leone, including three genomes from the initial Guinean introduction, for a total of 514 genomes^12,13,18^. This broader dataset allowed transmission chains to extend outside the district, placing the KGH cluster within the larger Sierra Leone epidemic and improving estimation of the shared parameters, including the substitution rate and estimated outbreak origin date, that constrained the reconstruction.

The inferred epidemic parameters were consistent with the known epidemiology and evolution of the West African epidemic. The *juniper*-inferred evolutionary rate was 1.16 x 10^-3^ substitutions per site per year (95% CrI 1.05 x 10^-3^ – 1.28 x 10^-3^, Figure 2C), consistent with published estimates for the 2013–2016 epidemic.^12,22^ The model placed the origin of the transmission chains represented in this dataset on 17 Feb 2014 (95% CrI 14 Jan – 05 Mar 2014; Figure 2B). Because the dataset included Guinean and non-Kenema Sierra Leonean sequences, this estimate reflects the origin of the sampled epidemic rather than the introduction of Ebola into Kenema District or the KGH workforce. The time-dependent reproductive number (R^t^) peaked at 3.29 (95% CrI: 2.22–4.03) on 29 May 2014 and did not decline below 1 over the duration of collected samples (Figure 2D).

**Figure 2.**
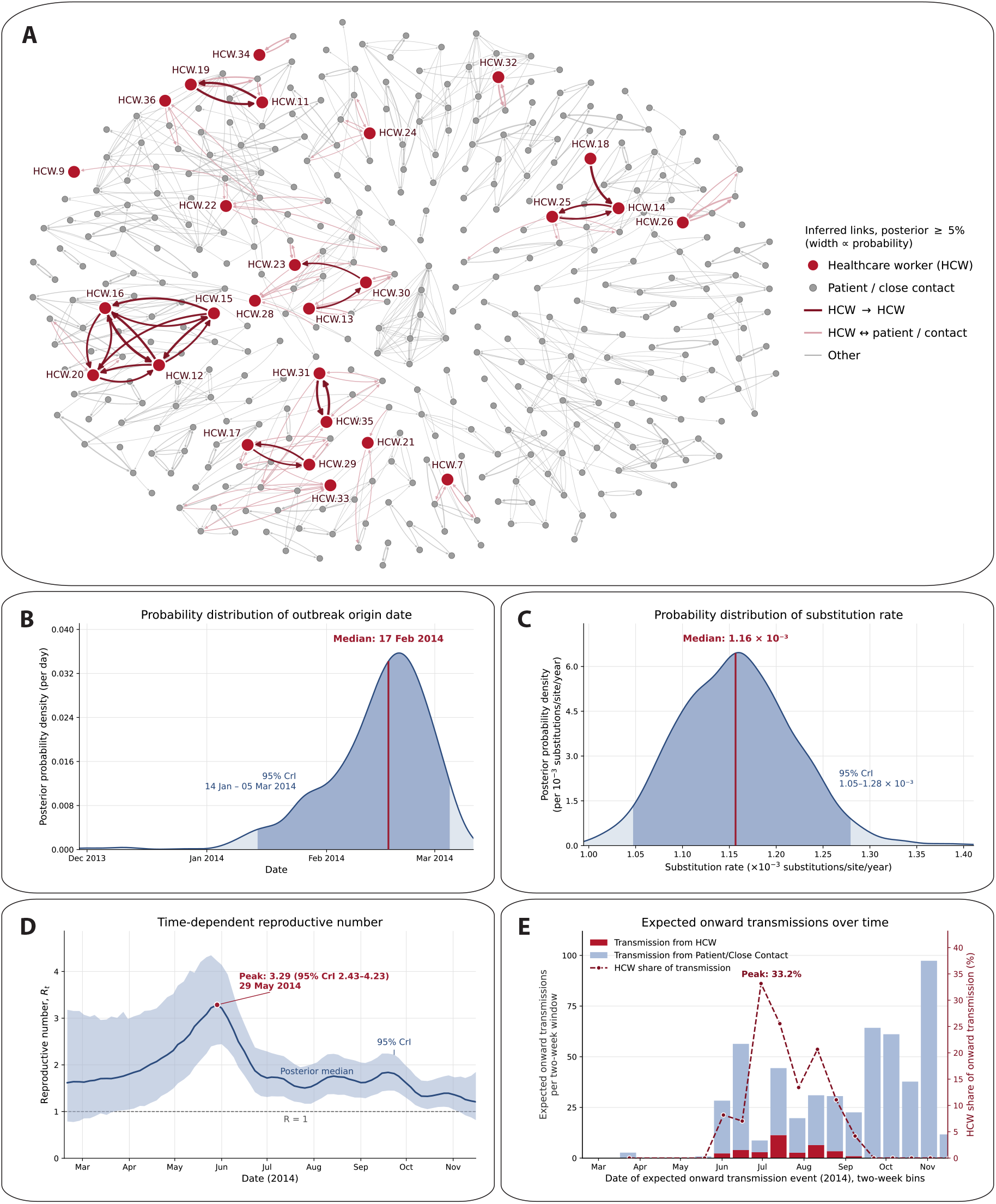
Transmission reconstruction of the Kenema Ebola outbreak with *juniper*. **(A)** Pairwise transmission network inferred across 514 sampled EVD cases using *juniper*. Nodes represent individual cases colored by status (red, healthcare worker; yellow, unknown patients/close contacts). Gray edges indicate inferred transmission links with posterior probability ≥ 5%, with edge width proportional to that probability; red edges highlight the 22 inferred healthcare worker-to-healthcare worker transmission links. HCW nodes are labeled. **(B)** Posterior distribution of the inferred outbreak start date (median 17 February 2014; 95% credible interval [CrI] 14 January–05 March 2014; effective sample size [ESS] = 980). **(C)** Posterior distribution of the substitution rate (median 1.16 x 10^-3^ substitutions/site/year; 95% CrI 1.05 x 10^-3^–1.28 x 10^-3^; ESS = 262). Dashed lines in (B)–(D) indicate posterior medians. **(D)** Time-dependent reproductive number, shown as the posterior median (solid line) with 95% CrI (shaded band); minimum ESS = 599. The dashed horizontal line marks R = 1. The inferred outbreak start (17 February 2014) precedes the plotted window. **(E)** Expected number of onward transmissions per two-week window, attributed to the infecting case and stratified by that case’s status. Stacked bars show offspring means averaged across posterior transmission trees, including transmissions to unsampled cases, for infectors classified as healthcare workers (dark red) or unknown (light blue). The dashed line indicates the percentage of expected transmissions in each window attributable to HCWs, plotted against the right axis.

Transmission inference revealed that healthcare workers formed a disproportionately interconnected network within the broader outbreak (Figure 2A). Of 800 links inferred with posterior probability ≥ 0.05, 22 links connected HCWs to other HCWs, compared with 3.6 expected when HCW labels were randomly permuted across nodes while holding the reconstructed network fixed. HCW-to-HCW transmission links were therefore enriched 6.1-fold over expectation (p < 0.0001, 10,000 permutations). Together, these findings indicate that once Ebola virus entered the healthcare workforce, transmission propagated substantially within that workforce rather than arising solely from repeated independent patient-to-HCW exposures.

HCW-driven transmission was concentrated in a short period during the early expansion of the outbreak at KGH. Among sampled cases in the reconstructed transmission network, HCWs accounted for less than 10% of expected onward transmissions in early June, peaking at 33.2% in the window beginning 23 Jun 2014, and less than 20% again by September (Figure 2E). Because sequencing coverage was substantially denser within Kenema District than elsewhere in the sampled region, these proportions describe the composition of the reconstructed network rather than the epidemic as a whole. Within that network, the HCW contribution rose and fell over a period of weeks rather than tracking the accumulation of cases, indicating that transmission through the healthcare workforce acted as a transient amplification pathway during the period of steepest local growth.

### Epidemiologic accounts situate inferred HCW transmission within the hospital

Our genomic analyses identified potential transmission relationships among healthcare workers but could not establish where exposures occurred or what clinical circumstances connected individual cases. We therefore integrated the genomic reconstruction with epidemiologic records and firsthand accounts from KGH staff, including one of the authors (M.M.), who worked at KGH during the outbreak. These accounts helped contextualize inferred links through specific wards, clinical procedures, and working relationships within the hospital (Figure 3).

**Figure 3.**
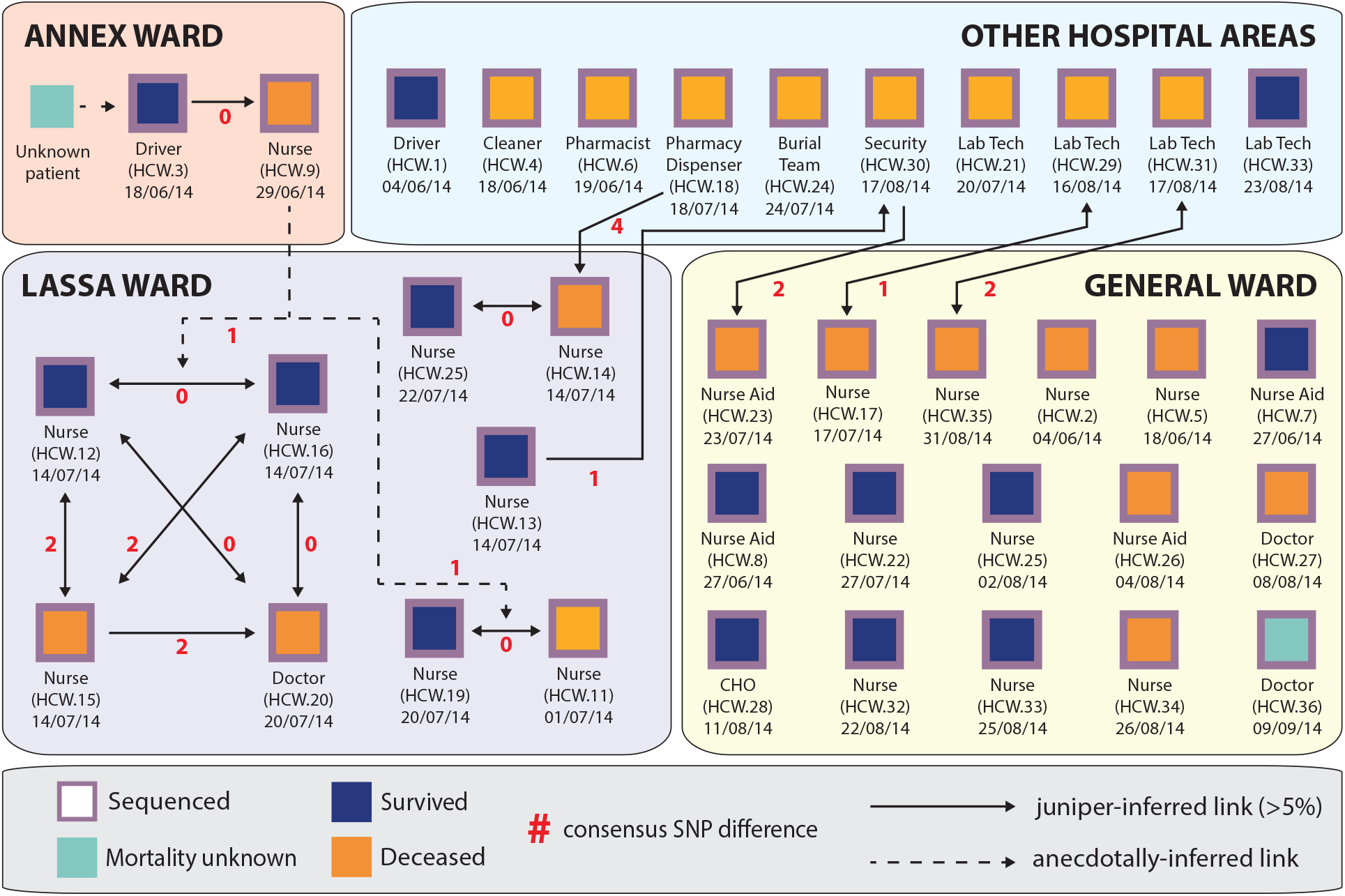
Reconstructed transmission relationships among healthcare workers at Kenema Government Hospital during the 2014 Ebola outbreak. Each box represents a HCW, labeled by anonymized case identifier, occupational role, and date of symptom onset or confirmed diagnosis. Cases are grouped by ward or hospital area (Annex Ward, Lassa Ward, General Ward, and Other Hospital Areas). These groupings reflect the hospital’s original ward structure; as the outbreak overwhelmed the Lassa Ward, Ebola care expanded throughout KGH, with the remaining wards converted into Ebola treatment units by mid-August 2014. Box color indicates clinical outcome (dark blue, survived; orange, deceased; teal, mortality unknown), and purple borders indicate availability of a viral consensus sequence. Solid arrows represent transmission links inferred by *juniper* transmission inference; bidirectional arrows indicate bidirectional inference in transmission direction. Dashed arrows indicate additional epidemiologically plausible links supported by KGH firsthand accounts but not inferred by *juniper*. Red numerals indicate pairwise consensus SNP differences between sequenced cases. The schematic integrates genomic reconstruction with epidemiologic accounts to illustrate potential transmission pathways across the hospital workforce; predicted links do not establish absolute direct transmission or the precise setting of exposures.

The earliest reconstructed HCW infections linked a hospital outreach driver with a nurse who cared for him. HCW.3, a driver for the KGH Lassa fever outreach team, had recently traveled to Koindu, where Ebola had circulated before its recognition in Kenema. The driver’s viral sequence was identical at the consensus level to that of an unidentified patient. HCW.3 was admitted to the Annex Ward on June 18, 2014, when consistent use of appropriate PPE had not yet been established in the ward. HCW.9, a nurse who cared for HCW.3, developed illness eleven days later. The two HCWs’ viral sequences were also identical at the consensus level. Together, the genomic similarity, documented care relationship, and timing of illness supported a transmission link between HCW.3 and HCW.9, although the sequence data alone could not establish direction.

As Ebola spread among KGH staff, caring for infected colleagues under severe resource constraints created additional opportunities for HCW-to-HCW exposure. When HCW.9, who was pregnant, deteriorated clinically, nurses HCW.11 and HCW.19 attempted a stillbirth delivery, a procedure routinely performed in Lassa fever care. Both nurses developed symptoms in the following weeks and subsequently tested positive for Ebola. Their viral consensus sequences were identical, and *juniper* inferred a potential transmission link between them. Their involvement in HCW.9’s care represented a plausible shared exposure, although the available evidence could not distinguish direct transmission between the nurses from infections acquired through a common source.

A further cluster subsequently emerged in the Lassa Ward, involving three nurses and a physician (Figure 3). HCW.12, HCW.15, and HCW.16 tested positive for Ebola on July 14, 2014, followed six days later by HCW.20, a physician on the ward. HCW.12, HCW.16, and HCW.20 shared identical viral consensus sequences, while HCW.15 differed by two consensus SNPs (Figure 1B–C). The *juniper* reconstruction identified multiple potential transmission links among these four HCWs (Figure 2A), consistent with the closely connected relationships depicted in Figure 3. Their shared clinical setting, closely spaced diagnoses, and genomic relationships support a cluster of epidemiologically linked infections within the Lassa Ward, although the precise sources and direction of transmission remain uncertain.

Transmission relationships also extended beyond HCWs providing routine bedside care with direct patient contact. HCW.18, a pharmacy dispenser, carried a viral consensus sequence differing by one SNP from that of HCW.3, the driver involved in the early Annex Ward exposure (Figure 1B–C). The *juniper* reconstruction identified a potential transmission linkage between HCW.18 and HCW.14, a Lassa Ward nurse, whose sequences differed by four consensus SNPs (Figures 2A, 3). HCW.14 and HCW.25, another Lassa Ward nurse, shared identical viral consensus sequences and were also linked in the transmission reconstruction (Figures 1B–C, 2A, 3). These relationships suggested connections between clinical staff and personnel working elsewhere in the hospital, although the precise exposure settings and transmission pathways remained uncertain. Other infected HCWs included HCW.24, a member of the hospital burial team, whose duties involved handling the bodies of deceased patients and presented distinct occupational exposure risks (Figure 3).

As Ebola patient admissions overwhelmed the Lassa Ward, KGH progressively converted its remaining wards into designated Ebola treatment areas, with routine hospital services for other illnesses effectively coming to a halt by late-August. Infections among HCWs continued amid this hospital-wide reorganization, and inferred transmission relationships involved staff across the broader hospital wards. The *juniper* reconstruction linked HCW.17, a General Ward nurse, with HCW.29, a laboratory technician, whose viral sequences differed by one consensus SNP, and HCW.31, another laboratory technician, with HCW.35, a General Ward nurse, whose sequences differed by two SNPs (Figures 1B–C, 2A, 3). The broader network also identified potential links involving HCW.30, a security officer, and HCW.13 and HCW.23, a Lassa Ward nurse and General Ward nursing aide, respectively (Figure 2A). Other HCW infections, including HCW.28, HCW.32, HCW.33, and HCW.36, occurred later in the outbreak but were not connected by specific inferred pairwise links (Figures 1A, 3). These findings suggest that infections among HCWs extended across multiple occupational groups and continued beyond the initial Lassa Ward cluster, although the available evidence could not determine whether individual infections arose through occupational exposure, contact outside the hospital, or separate introductions into the workforce.

Together, the genomic and epidemiologic evidence supports transmission among healthcare workers across multiple hospital settings. Where sequence data were unavailable, we inferred links by combining epidemiologic accounts with the genomic comparisons among sequenced cases in the proposed chain; these links carry greater uncertainty. Even with its limitations the combined reconstruction supports a coherent pattern: an early exposure near the hospital’s periphery, movement through the Annex and General Wards, further amplification during care of infected colleagues and within the Lassa Ward, and additional infections among staff outside direct bedside care before a later, distinct HCW cluster emerged.

## Discussion

Our study of KGH during the 2014 Ebola outbreak highlights a critical vulnerability in outbreak response, showing how HCWs are not only at risk of infection from the patients they treat but can themselves become an interconnected network through which infection spreads and amplifies. Within the reconstructed network, we identified 22 potential HCW-to-HCW transmission links, compared with 3.6 expected if HCW status were randomly assigned across cases, representing a 6.1-fold enrichment. Among cases with inferred infection dates in the two-week window beginning June 23, 2014, HCWs accounted for 33.2% of expected onward transmissions, compared with under 10% for cases infected in early June. Earlier epidemiologic investigations at KGH documented care of infected colleagues and other HCW-to-HCW exposures as recurrent features of the outbreak. Our analysis builds on those observations by reconstructing probable transmission relationships and showing how the HCW contribution changed over the course of the outbreak^6^.

Genomic epidemiology can reveal patterns of viral spread, but understanding how transmission unfolds within a health system also requires examining the human relationships and circumstances behind those patterns. At KGH, expanded genomic sampling identified closely related HCW clusters, but classical genomic analyses could not resolve individual transmission pathways. We used *juniper* transmission inference to estimate posterior probability for alternative transmission links while explicitly accounting for unsampled cases. Epidemiologic records and firsthand accounts then situated inferred links within specific wards, clinical events, and working relationships, including care of infected colleagues under severe resource constraints. By integrating these approaches, we could better understand the circumstances and vulnerabilities that may have enabled transmission within the healthcare workforce, while retaining uncertainty about individual sources of infection.

KGH illustrates the danger of an outbreak outpacing the resources needed to provide safe care. Despite longstanding experience with viral hemorrhagic fever, KGH could not compensate for shortages of protective supplies, personnel, and isolation capacity as Ebola admissions increased.^6,15^ HCW infections peaked before or during the expansion of external support, although the timing alone cannot establish the effects of individual interventions (Figure 4). Preparedness plans should therefore equip frontline hospitals with protective supplies in advance and provide for rapid expansion of staffing and isolation capacity at the first signs of an outbreak.

**Figure 4.**
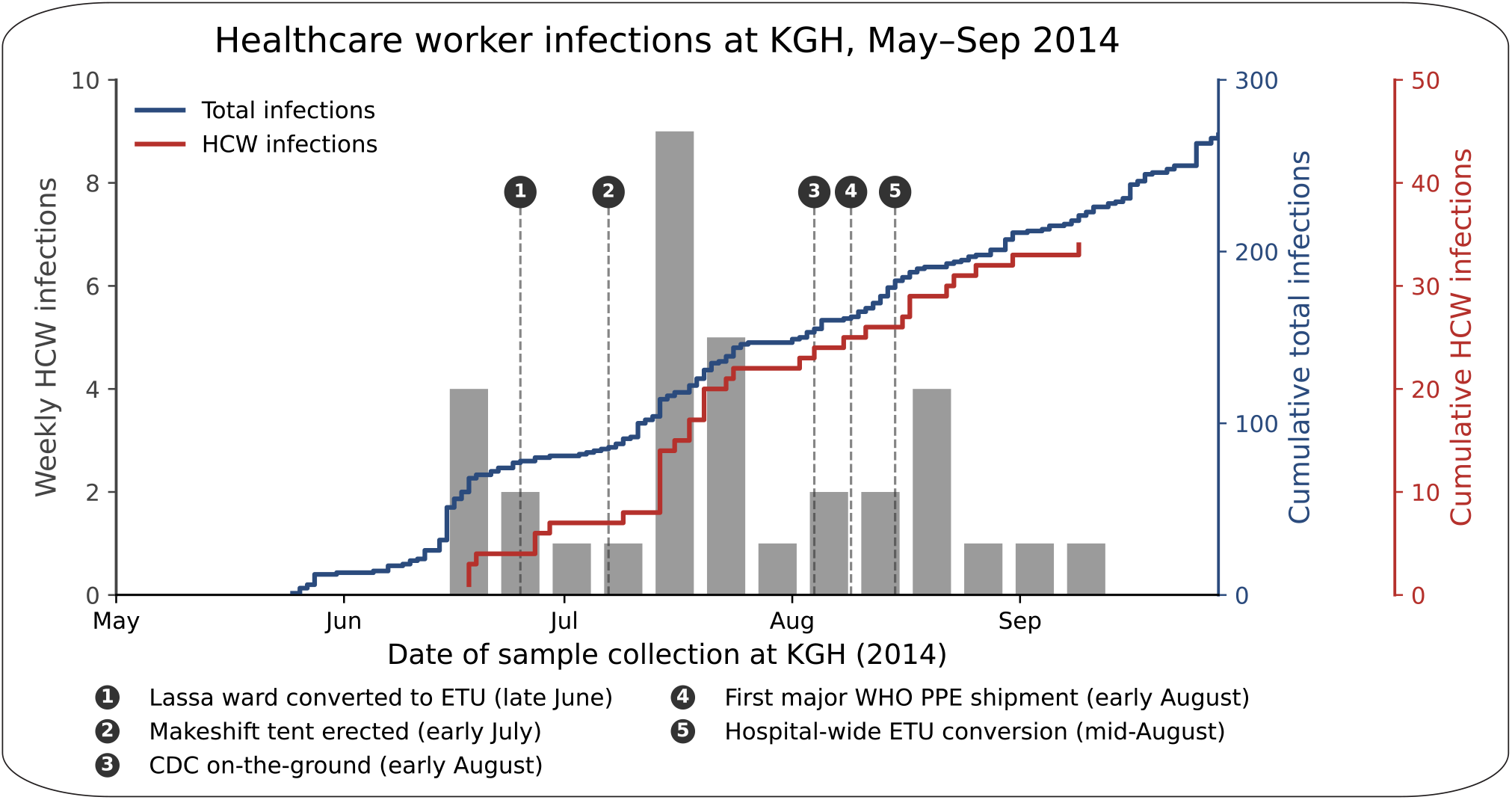
Timing of Ebola infections among KGH healthcare workers and arrival of external response support, 2014. The gray bars show weekly HCW infection counts at KGH, plotted on the left axis. The blue line shows cumulative total Ebola infections sequenced at KGH over the same period, plotted on the right axis. The red line shows cumulative HCW Ebola infections sequenced at KGH over the same period, plotted on the right axis. Vertical dashed lines mark key milestones in the expansion of KGH’s Ebola response: (1) conversion of the Lassa ward into the country’s first Ebola Treatment Unit (late June); (2) erection of a makeshift treatment tent (early July); (3) arrival of CDC personnel on the ground (early August); (4) arrival of the first major WHO PPE shipment (early August); and (5) conversion of the hospital’s remaining wards into Ebola treatment units (mid-August). The highest weekly burden of HCW infections occurred before or during the early phase of this response scale-up, highlighting the vulnerability of frontline staff during the interval between Ebola’s arrival at KGH and the availability of sustained protective infrastructure.

Outbreak response must not only protect healthcare workers during patient care but also prevent infection from spreading among colleagues. Staff with repeated patient exposures continue to interact through handoffs, shared workspaces, meals, transportation, and care of sick colleagues. Without early detection and adequate protection, one infection can spread among colleagues and weaken the very team needed to care for patients. Healthcare workers should therefore be a priority surveillance population, with rapid access to testing, active investigation of clusters among clinical and support staff, and infection-control practices that explicitly consider transmission between colleagues.

Although we identified broader patterns of transmission within the healthcare workforce, our study has several limitations, particularly in resolving individual transmission links. We analyzed genomes from approximately 47% of confirmed EVD cases in Kenema District and 44% of infected HCWs, leaving unsampled intermediates plausible for many links. Uneven sequencing coverage also means that the inferred HCW contribution to onward transmission reflects the reconstructed network rather than the epidemic as a whole. Although *juniper* models unsampled cases, its transmission assignments remain probabilistic and depend on the model’s assumptions^19^. Close genomic relationships alone cannot establish direct transmission or exclude shared patient exposures and community acquisition.^25^ We also could not account systematically for social or personal interactions among healthcare workers outside the hospital, and some transmission attributed to workplace relationships may instead have occurred through contact between colleagues in the community or while caring for one another outside formal clinical settings. The staff recollections we used in the study may be incomplete or inaccurate, even when case records support the reported timing. We relied on those accounts to help interpret the inferred links, not to confirm them independently.

The 2026 Bundibugyo virus outbreak in the Democratic Republic of the Congo underscores both the continuing vulnerability of HCWs and the importance of genomic epidemiology in revealing transmission risks. HCWs were among the earliest recognized fatalities, and no vaccine or specific treatment is currently approved for Bundibugyo virus disease^26,27^. Continued genomic epidemiologic investigations, integrated with local epidemiologic knowledge and firsthand accounts, may reveal other vulnerabilities that would otherwise remain hidden. Protecting healthcare workers requires looking beyond individual exposures to safeguard the interconnected teams on which outbreak response depends.

## Methods

### Patient sample collection

Research involving human subjects at the Kenema Government Hospital was approved by the Sierra Leone Ethics and Scientific Review Committee, Tulane University and Harvard University Institutional Review Boards (IRBs) in accordance with National Institutes of Health guidelines. All patients enrolled through 25 May 2014 provided informed consent and were enrolled as having suspected Lassa fever prior to the first diagnosis of an Ebola case in Sierra Leone. Because of the Ebola outbreak, a waiver of written consent was granted for testing samples obtained from patients who enrolled after 25 May. Scientists wishing to obtain a key linking the current case designations of deidentified patients to previously published deidentified patient identifiers should contact the authors.

To gain a deeper understanding of transmission we first identified additional samples stored at the KGH biobank for viral sequencing. From June through November 2014, 591 cases of EVD originated in Kenema District, including 79 (15%) from HCWs. We identified 14 HCW sequences previously sequenced and generated 22 new sequences from known HCWs for a total of 36 sequences, representing approximately half of infected HCWs in Kenema District^12,18^. We also generated 42 new Ebola sequences from the general population of Kenema District collected around the same time as the HCWs. Sequencing of these samples occurred at the Kenema Government Hospital’s sequencing core and the Broad Institute. Combined with sequences obtained in our earlier studies this brings the number of sequences from Kenema District to 280, 47% of the total EVD cases reported during this period.

### Sample Processing

Serum/plasma from whole blood was prepared according to the method described in (Matranga *et al*., 2014) and, used to extract Ebola RNA using QIAamp Viral RNA Minikit (Qiagen) according to manufacturer’s instructions^28^. Briefly, 140 µl of serum/plasma were inactivated by AVL and ethanol treatment. Purification of extracted RNA was completed by washing the spin column twice using antiviral wash buffers (AW1& AW2). Purified RNA was then eluted in 60 µl of elution buffer in a 1.5ml eppendorf tube and kept in a -20°C freezer while in processing and in a -80°C freezer for a longer period during shipments.

We utilized Sybr Green RNA-to-Ct 1-Step qRT-PCR assay (Life Technologies) for absolute quantification of Ebola virus genome in real-time. We used Kulesh primer set (Forward and Reverse) to amplify region within the NP gene according to protocol previously described in (Drosten *et al*., 2002) on Roche LightCycler 96 with some modification^29^. Briefly, kulesh primers were used at a loading concentration of 10 µM and a reaction volume of 10 µl containing 3µl of RNA and 7µl of master mix was used and, each sample was run in triplicate.

### Viral Sequencing

Towards constructing sequencing libraries, samples were enriched by both 18s depletion and turbo DNase treatment and, cDNA was synthesized in batches as previously described (Adiconis *et al*., 2013 was referenced in Matranga *et al*., 2014 for 18s depletion (26))^28,30^. Sequencing libraries were constructed using Illumina Nextera library construction kits according to the methods of Matranga *et al*.^28^. Complete sequencing libraries were cleaned up using Ampure DNA SPRI and, quantified by both KAPA qPCR and BioAnalyzer using the manufacturer’s instructions. Pooled libraries were paired-end sequenced at a loading concentration of 12 pM for 101 cycles using Illumina V2 kit on Illumina MiSeq platform.

### Genomic Analysis

Consensus genomes were aligned to the Ebola virus reference KJ660346.2, following the parameters from *Gire et al*.. Alignments were screened for clustered same-direction transitions before analysis, as ADAR-mediated editing can introduce runs of T-to-C substitutions within a short genomic interval that do not represent independently accumulated mutations as we previously reported in *Park et al*.^18^. Genomic sites were masked where 5 or more same-direction T-to-C transitions occurred within a consecutive 200bp window. One such cluster met this criterion: eight T-to-C substitutions between positions 5515 and 5560 in HCW.11 and HCW.19. These sites were masked in all downstream analyses, including phylogenetic reconstruction, haplotype network construction, and transmission inference.

Maximum-likelihood phylogenies were inferred in IQ-TREE, with the best-fitting nucleotide substitution model selected using ModelFinder and branch lengths estimated in substitutions per site^21^. Trees were visualized in FigTree v1.4.4, with patient-only clades collapsed for display^31^. Pairwise SNP distances among the 35 HCW consensus genomes were computed directly from the masked multiple-sequence alignment, excluding sites with ambiguous calls in either sequence. The haplotype network was constructed from the 35 HCW consensus genomes: sequences with identical genotypes across all variable sites were collapsed into single haplotypes. Each edge is annotated with the specific genomic positions differentiating the haplotypes it connects. Because the network is a minimum spanning tree, a single topology is shown where several equally parsimonious connections may exist; alternative resolutions are not displayed.

Analyses and figures were generated in Python v.3.12.4.

### Transmission Reconstruction Analysis

Transmission histories were reconstructed using *juniper*, a Bayesian framework that jointly infers the phylogeny and the transmission network from consensus genome sequences, sampling times, and case metadata, while explicitly modeling unsampled cases^19^.

The model was fit to 521 Ebola virus consensus genomes with associated collection dates. 7 genomes were excluded due to a set sample cutoff date of Nov 16, 2014, resulting in a total of 514 genomes included in the *juniper* analysis. Priors were specified as a multivariate normal distribution on the log-transformed parameter vector (log λ^T^, log ψ, log t^root^, log μ), with mean (2.904165, 0.60158, 0, ™6.9077551) and identity covariance matrix, giving independent log-normal priors with unit standard deviation on the log scale. This corresponds to prior medians of 18.25 year^-1^ for the transmission rate λ^T^, 1.8 year^-1^ for the sampling rate ψ, 1 year for the root time t^root^, and 1.0 x 10^-3^ substitutions per site per year for the substitution rate μ, each with a 95% prior interval spanning roughly a seven-fold range above and below the median (for example, 1.41 x 10^-4^ to 7.10 x 10^-3^ for μ). Inference was run for 10,000 MCMC iterations, with the first 20% discarded as burn-in. Convergence was assessed by effective sample size (ESS), which exceeded 200 for all reported parameters (outbreak start date, ESS = 980; substitution rate, ESS = 262; time-dependent reproductive number, minimum ESS = 599).

Posterior estimates were summarized as medians with 95% credible intervals. The substitution rate was estimated at 1.16 x 10^-3^ substitutions per site per year (95% CrI 1.05 x 10^-3^ – 1.28 x 10^-3^; Figure 2C) and the outbreak start date at 17 February 2014 (95% CrI 14 January–05 March 2014; Figure 2B). The time-dependent reproductive number, R = λ/(γ + ψ), is reported as the posterior median with 95% credible interval (Figure 2D).

Pairwise transmission probabilities were extracted as the posterior frequency with which each ordered pair appeared as a direct transmission link across sampled trees. The network in Figure 2A displays edges with posterior probability ≥ 0.05, with edge width proportional to probability; links inferred between two healthcare workers are highlighted separately. To produce Figure 2E, each inferred transmission event was assigned to the two-week window containing the infector’s infection date and classified by the infector’s status (healthcare worker, close contact, or unknown), and the resulting offspring means were averaged across posterior trees, including transmissions to unsampled cases.

### Anecdotal contact tracing

Kenema Government Hospital housed Sierra Leone’s national Lassa fever laboratory and was, throughout the epidemic, the only facility in the country with established infrastructure for viral hemorrhagic fever diagnostics, surveillance, and clinical care^15^. The KGH viral hemorrhagic fever team operated on behalf of and in collaboration with the Sierra Leone Ministry of Health and comprised clinical, laboratory, and outreach staff, several of whom are authors on this study.

Contextual information on ward assignments, clinical procedures, and working relationships among KGH staff was obtained from firsthand accounts provided by team members who worked at KGH throughout the 2014 outbreak, including M.M., who led the KGH laboratory; L.K., who led the KGH outreach team; A.G., director of the viral hemorrhagic fever laboratory; and other hospital staff. Reported details were corroborated where possible against laboratory case logs and dates of symptom onset or confirmed diagnosis held at KGH. Details that could not be independently verified against these sources are identified as such.

Accounts were used to situate transmission relationships already inferred from the genomic data rather than to generate candidate links independently. For each pair of cases connected in the reconstruction, we asked whether the two individuals had occupied a common ward, participated in a shared clinical procedure, or maintained a professional or personal relationship that would have likely brought them into contact or exposure during the relevant interval. Where sequence data were unavailable for an individual named in an account, that case was positioned in the reconstruction by triangulating the reported epidemiologic contacts against the SNP distances separating the nearest sequenced cases on either side. Links inferred in this way carry greater uncertainty than those supported directly by sequence comparison and are indicated in Figure 3.

Case identifiers (HCW.1–HCW.36) were assigned to de-identify individual healthcare workers, and the mapping between identifiers and genomic sequence labels is held in a restricted key available to qualified researchers on request. Roles are reported at the level of occupational category (nurse, physician, laboratory technician, driver, pharmacy dispenser, burial team member) rather than by individual post.

## Data Availability

Sequence data are available at NCBI (NCBI BioGroup: PRJNA257197). All other data produced in the present study are available upon reasonable request to the authors.

## Acknowledgements

Augustine Goba, a co-author and longtime leader of the Kenema Government Hospital Lassa fever program, passed away before publication of this work. We are deeply grateful for his contributions to this study and to the care of patients and healthcare workers in Sierra Leone.

We dedicate this work to the healthcare workers of Kenema Government Hospital who served during the 2014 Ebola outbreak, to those who lost their lives, and to their families. We also dedicate it to healthcare workers everywhere who place themselves at risk during outbreaks, and to the families who support and share in that sacrifice.

We thank the Sierra Leone Ministry of Health and Sanitation, the Kenema District Health Management Team, Kenema Government Hospital, and other partners for their efforts in outbreak response.

